# Ophthalmic artery stenosis on three-dimensional rotational angiography: interrater agreement, prevalence, and risk factors

**DOI:** 10.1101/2023.11.08.23298282

**Authors:** William K Diprose, Michael T. M. Wang, Joseph Reidy, Alice Ma, James Brodie, Brendan Steinfort

## Abstract

**Background:** There is emerging interest in ophthalmic artery stenosis angioplasty for the treatment of age-related macular degeneration. Three-dimensional rotational angiography could be used intraoperatively to determine the presence and severity of ophthalmic artery stenosis. In patients who had undergone three-dimensional rotational angiography of the internal carotid artery, we aimed to assess the interrater agreement, prevalence, and risk factors for ophthalmic artery stenosis.

**Methods:** Consecutive patients from two centers who had undergone diagnostic cerebral angiography with three-dimensional rotational angiography of the internal carotid arteries were enrolled in this study. Three-dimensional rotational angiograms were independently double read for the presence of ophthalmic artery stenosis, as defined as an ostial narrowing of at least 50% when compared to the more distal ‘normal’ ophthalmic artery. Interrater agreement for the evaluation of ophthalmic artery stenosis was assessed with the Cohen’s kappa coefficient. Univariate and multivariable logistic regression were used to identify potential predictors of OA stenosis.

**Results:** 302 patients (97 men; mean±SD 57.6±13.4 years) were included in the analysis. Cohen’s kappa coefficient (95% CI) was 0.877 (0.798–0.956). Ophthalmic artery stenosis was present in 45 patients (14.9%). Multiple logistic regression demonstrated that female sex was independently associated with a higher odds of ophthalmic artery stenosis (odds ratio [OR]=2.70, 95% confidence interval [CI] 1.18-6.09, P=0.02). Smoking was a significant risk factor for ophthalmic artery stenosis (OR=2.11, 95% CI 1.10-4.06, P=0.03).

**Conclusion:** The evaluation of ophthalmic artery stenosis on three-dimensional rotational angiography had excellent interrater agreement. Ophthalmic artery stenosis was common and was associated with smoking and female sex.

## INTRODUCTION

Ophthalmic artery stenosis is a poorly understood clinical entity that has historically been described as a rare cause of amaurosis fugax.^1–3^ However, there is growing interest in the role of impaired ophthalmic artery blood flow in the pathogenesis of age-related macular degeneration,^4,5^ with a small case series of five patients demonstrating improved vision following balloon angioplasty of the ophthalmic artery ostium.^6^ In this series, preoperative 3T magnetic resonance angiography was used to identify compromised blood flow in the ophthalmic artery. Intraoperatively, three-dimensional rotational angiography was used to confirm ophthalmic artery stenosis amenable to balloon angioplasty. No agreed upon definition of ophthalmic artery stenosis exists, and the interrater agreement of the assessment of ophthalmic artery stenosis on three-dimensional rotational is unknown. We aimed to assess the interrater agreement, prevalence, and risk factors for ophthalmic artery stenosis in patients who had undergone three-dimensional rotational of the internal carotid artery.

## METHODS

### Study design

The study is reported according to the Guidelines for Reporting Reliability and Agreement Studies recommendations.^7^ The data that support the findings of this study are available from the corresponding author upon reasonable request. This study was approved by the regional ethics committee (2022/ETH01961) and informed consent was not required.

Consecutive patients from two centers (Royal North Shore Hospital and Westmead Hospital, NSW, Australia) who had undergone diagnostic cerebral angiography with three-dimensional rotational angiography of the internal carotid arteries were retrospectively enrolled in this study. Patients were consecutively enrolled until at least 40 cases of ophthalmic artery stenosis were identified, allowing for up to four covariates to be included in a multivariable analysis, with the number of events per variable being 10.^8^ Patients were excluded if they had no ophthalmic artery arising from the internal carotid artery (e.g., those with aberrant ophthalmic arteries arising from the middle meningeal artery), paraophthalmic aneurysms, or significant ophthalmic carotid segment atherosclerotic disease. The source three-dimensional rotational angiograms were independently double read by a neurointerventionalist (B.S, 19 years’ experience) and a neurointerventional fellow (W.K.D, 1 year experience) for the presence of ophthalmic artery stenosis. As no published standard for classifying or grading of ophthalmic artery stenosis exists, a modified North American Symptomatic Carotid Endarterectomy Trial criteria for arterial stenosis was used.^9^ The diameter of the ophthalmic artery ostium was compared with the more distal ‘normal’ ophthalmic artery. If there was narrowing of ≥50% (i.e., at least moderate stenosis), this was classified as ophthalmic artery stenosis (Figure 1).^10^ Disagreements were resolved by a third neurointerventionalist (A.M, 5 years’ experience). Potential risk factors for ophthalmic artery stenosis, including age, sex, smoking history (current or ex-smoker), hypertension, diabetes, and cardiovascular disease were extracted from the electronic health record.

**Figure 1.**
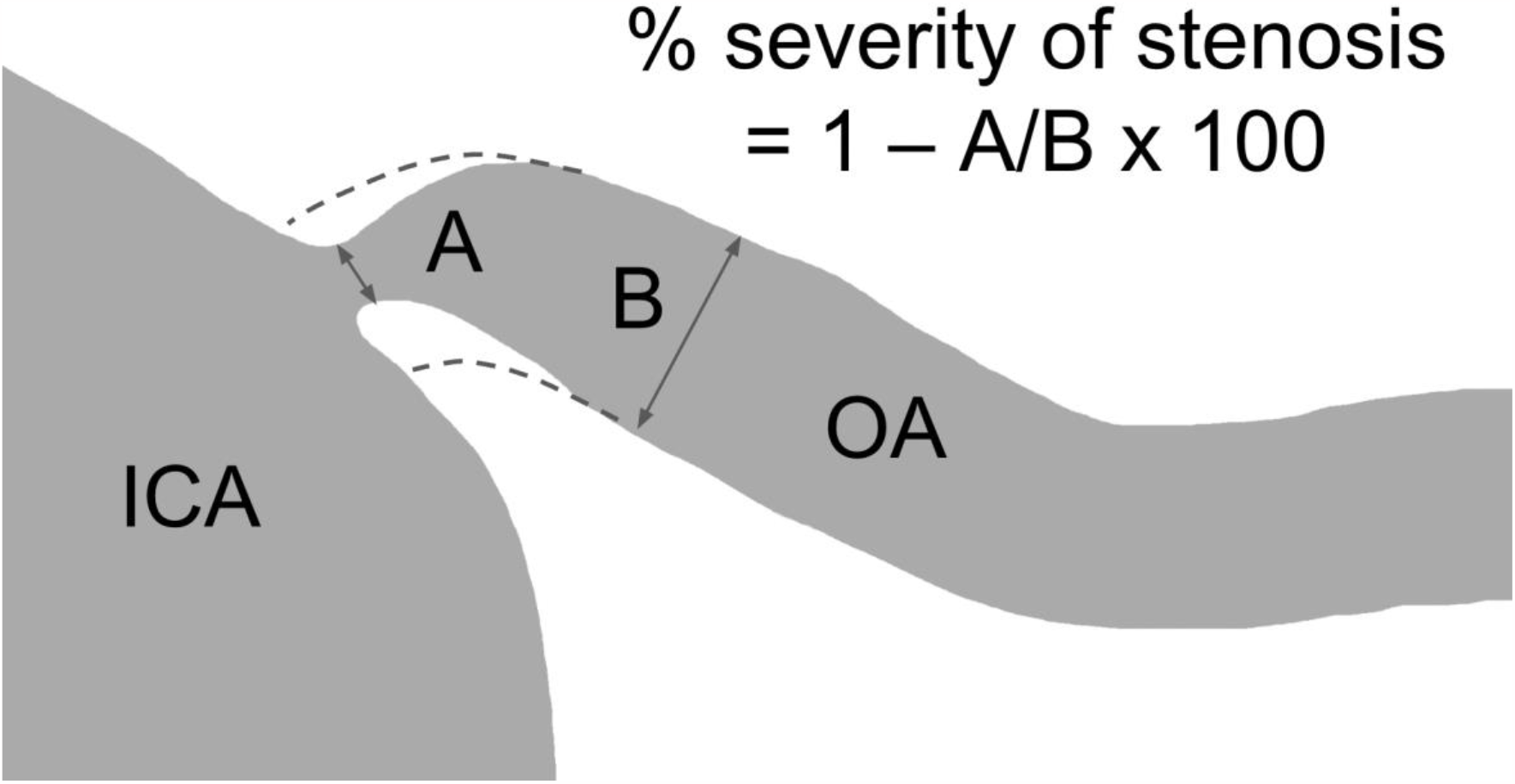
Modified North American Symptomatic Carotid Endarterectomy Trial criteria for the diagnosis of ophthalmic artery stenosis. ICA = internal carotid artery; OA = ophthalmic artery.

### Statistics

Statistical analysis was conducted with Graph Pad Prism version 8.01 (California, USA) and IBM SPSS version 24 (New York, USA). Interrater agreement for the evaluation of ophthalmic artery stenosis was assessed with the Cohen’s kappa coefficient. Preliminary univariate logistic regression was used to identify potential predictors of ophthalmic artery stenosis. Multivariable logistic regression for predictors of ophthalmic artery stenosis was then conducted, incorporating variables with a univariate association threshold of p<0.15.

The number of variables used in the multivariate regression analysis was limited to the number of diagnosed participants divided by 10, to avoid overfitting. All tests were two tailed, and p<0.05 was considered significant. Data are presented as mean±SD, or number of participants (% of participants) unless otherwise stated.

## RESULTS

There were 302 patients (97 men; mean±SD 57.6±13.4 years) included in the analysis (Table 1). The Cohen’s kappa coefficient (95% CI) for interrater agreement for the evaluation of ophthalmic artery stenosis was 0.877 (0.798–0.956). Ophthalmic artery stenosis was present in 45 patients (14.9%), including 24 (7.9%) patients with right-sided disease, 17 (5.6%) patients with left-sided disease, and 4 (1.3%) patients with bilateral disease. Figure 2 demonstrates examples of normal and stenotic ophthalmic arteries.

**Table 1.** Patient characteristics.

|  | Value (n=302) |
| --- | --- |
| **Demographics** |  |
| Age, years, mean±SD | 57.6±13.4 |
| Female sex, n (%) | 205 (67.9%) |
| **Indication for three-dimensional rotational angiography** |  |
| Aneurysmal subarachnoid hemorrhage | 125 (41.4%) |
| Dural arteriovenous malformation | 5 (1.7%) |
| Intracranial vasculopathy | 6 (2.0%) |
| Nonaneurysmal subarachnoid hemorrhage | 23 (7.6%) |
| Ruptured arteriovenous malformation | 4 (1.3%) |
| Unruptured intracranial aneurysm | 120 (39.7%) |
| Unruptured arteriovenous malformation | 8 (2.6%) |
| Other indication | 11 (3.6%) |
| **Ophthalmic artery stenosis, n (%)** |  |
| Right | 24 (7.9%) |
| Left | 17 (5.6%) |
| Bilateral | 4 (1.3%) |
| **Comorbidities, n (%)** |  |
| Hypertension | 141 (46.7%) |
| Diabetes | 35 (11.6%) |
| Smoking | 127 (42.1%) |
| Coronary artery disease | 26 (8.6%) |

**Figure 2.**
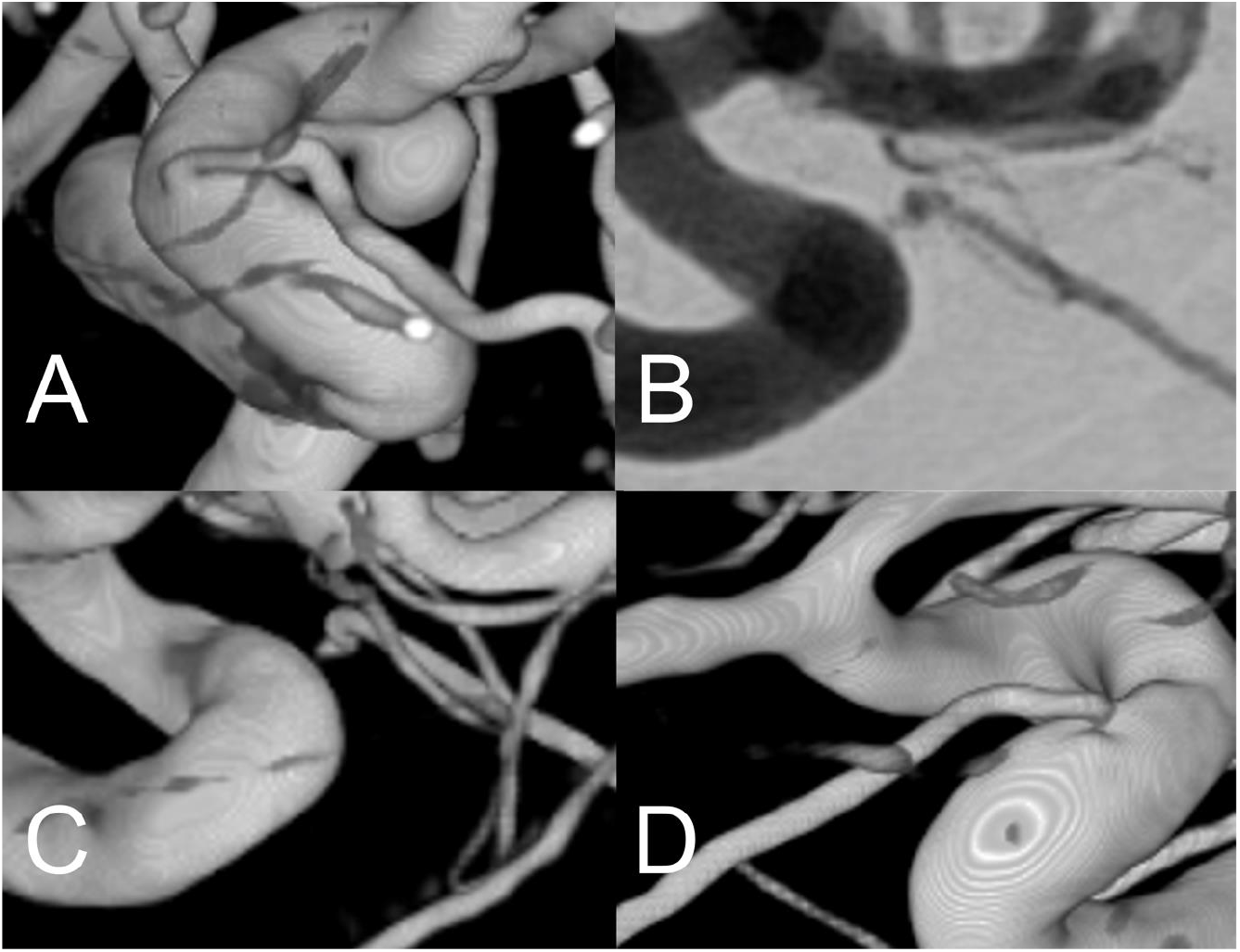
Three-dimensional rotational angiogram demonstrating a normal ophthalmic artery (A), and digital subtraction angiography (B) and three-dimensional rotational angiograms (C and D) demonstrating ophthalmic artery stenosis.

Unadjusted univariate and multivariable-adjusted odds ratios for the presence of ophthalmic artery stenosis by demographic and clinical factors are presented in Table 2. Multiple logistic regression demonstrated that female sex was independently associated with a higher odds of ophthalmic artery stenosis (odds ratio [OR]=2.70, 95% confidence interval [CI] 1.18-6.09, P=0.02). Smoking was a significant risk factor for ophthalmic artery stenosis (OR=2.11, 95% CI 1.10-4.06, P=0.03).

**Table 2.** Logistic regression odds ratio for the presence of ophthalmic artery stenosis by demographic and clinical factors.

| Characteristic | Unadjusted univariate logistic regression |  | Multivariable-adjusted logistic regression |  |
| --- | --- | --- | --- | --- |
|  | OR (95% CI) | P-value | OR (95% CI) | P-value |
| Age (per year) | 1.02 (1.00-1.05) | 0.10 | 1.02 (1.00-1.05) | 0.12 |
| Female sex | 2.44 (1.10-5.56) | 0.03 | 2.70 (1.18-6.09) | 0.02 |
| Hypertension | 0.81 (0.43-1.53) | 0.52 | - | - |
| Diabetes | 0.95 (0.35-2.58) | 0.91 | - | - |
| Smoking | 1.90 (1.00-3.59) | 0.05 | 2.11 (1.10-4.06) | 0.03 |
| Coronary artery disease | 1.82 (0.69-4.82) | 0.23 | - | - |
| Aneurysmal subarachnoid hemorrhage | 0.90 (0.48-1.69) | 0.73 | - | - |

## DISCUSSION

To the best of our knowledge, this is the first study to investigate the interrater agreement, prevalence, and risk factors of ophthalmic artery stenosis with three-dimensional rotational angiography. We found that the assessment of ophthalmic artery stenosis on three-dimensional rotational angiography had excellent interrater agreement with a Cohen’s kappa coefficient (95% CI) of 0.877 (0.798–0.956). Ophthalmic artery stenosis, as defined as an ostial narrowing of at least 50% when compared to the more distal ‘normal’ ophthalmic artery, occurred in 15% of our study cohort. Cigarette smoking and female sex were independently associated with a greater odds of ophthalmic artery stenosis.

To date, there have been limited studies investigating the clinical and radiologic features of ophthalmic artery stenosis. Ophthalmic artery stenosis was first described in 2/287 patients who underwent diagnostic cerebral angiography for the evaluation of carotid artery disease,^11^ and has subsequently been reported as a rare cause of amaurosis fugax.^1–3^ More recently, in a study of 101 patients who were treated with intra-arterial thrombolysis for acute non-arteritic central retinal artery occlusion, 38% had ophthalmic artery stenosis.^10^ When patients with mild stenosis (<50% narrowing) were excluded, only 18 patients (18%) had at least moderate (≥50% narrowing) ophthalmic artery stenosis, which is similar to our findings.

However, the authors used digital subtraction angiography and three-dimensional magnetic resonance contrast-enhanced angiography to diagnose and classify the severity of ophthalmic artery stenosis. Ophthalmic artery stenosis (and occlusion) has also been recently described as a complication of intra-arterial chemotherapy for childhood retinoblastoma.^12^

There is growing interest in the role of impaired ophthalmic artery blood flow in the pathogenesis of age-related macular degeneration,^4,5^ which is the leading cause of blindness among the elderly in the developed world and has limited treatment options.^13^ Smaller ophthalmic artery diameters and reduced ophthalmic artery volumetric flow rates were demonstrated with 7T non-contrast magnetic resonance angiography in patients with age-related macular degeneration when compared to healthy controls.^14^ The authors postulated that reduced choroidal perfusion could contribute to the development of age-related macular degeneration, but the study was unable to determine the direction of causality.

In a small series of five patients with late-stage age-related macular degeneration, balloon angioplasty of the ophthalmic artery ostium resulted in improvement in vision,^6^ suggesting a potential causal association between ophthalmic artery stenosis and visual impairment from age-related macular degeneration. Mean Snellen visual acuity was 20/710 pre-operatively and 20/406 six months postoperatively. In this series, preoperative 3T magnetic resonance angiography was used to identify compromised blood flow in the ophthalmic artery.

Intraoperatively, three-dimensional rotational angiography was used to confirm ophthalmic artery stenosis amenable to balloon angioplasty. The findings of the current study suggest that three-dimensional rotational angiography is a reasonable technique for diagnosing at least moderate ophthalmic artery stenosis prior to balloon angioplasty in future clinical trials.

The causes of ophthalmic artery stenosis are unknown. Aneurysmal subarachnoid hemorrhage was not associated with ophthalmic artery stenosis, suggesting that the high prevalence of ophthalmic artery stenosis was not likely explained by post-subarachnoid hemorrhage vasospasm. In contrast, cigarette smoking was associated with a two-fold risk of ophthalmic artery stenosis. Smoking-induced atherogenesis is well-described in the extracranial internal carotid arteries.^15^ However, the ophthalmic artery usually arises from the intracranial internal carotid artery,^16^ and the relationship between smoking and atherosclerosis of the intracranial internal carotid arteries is less well understood. For example, an early study demonstrated that duration of smoking was associated with atherosclerotic disease of the intracranial internal carotid arteries in both men and women.^17^ In a more recent population-based study of 2495 participants, smoking was associated with atherosclerotic disease of the intracranial internal carotid arteries in men, but not in women.^18^

Female sex was associated with 2.7 times increased odds of ophthalmic artery stenosis in the current study. This is an interesting finding, as men are more likely to have high-grade extracranial carotid stenosis,^19^ and there is no difference in the prevalence of intracranial atherosclerotic disease between the sexes.^20^ However, females have a higher risk of other non-atherosclerotic extracranial and intracranial vasculopathies, such as intracranial aneurysm disease,^21^ moyamoya disease,^22^ and fibromuscular dysplasia,^23^ raising the possibility that ophthalmic artery stenosis has a non-atherosclerotic pathology. On the other hand, patients with central retinal artery occlusion and ophthalmic artery stenosis were more likely to have a higher degree of stenosis and a more severe plaque morphology in the ipsilateral carotid artery, suggesting a shared atherosclerotic pathology.^10^

A causal relationship between ophthalmic artery stenosis and age-related macular degeneration has not yet been demonstrated. Age is the most important risk factor for age-related macular degeneration.^24,25^ In our multivariable logistic regression analysis, there was a trend towards older age and higher odds of ophthalmic artery stenosis, but this was not statistically significant. Because the majority (81%) of three-dimensional rotational angiograms were performed for the evaluation of ruptured or unruptured intracranial aneurysms, the mean age and standard deviation was not representative of the general population, which may have limited our ability to detect an association between age and ophthalmic artery stenosis.

Smoking is the most consistently reported modifiable risk factor for age-related macular degeneration,^26,27^ and female sex has been shown to be a risk factor for age-related macular degeneration in some studies.^13,28^ Smoking and female sex were both associated with ophthalmic artery stenosis in the current study, suggesting that age-related macular degeneration and ophthalmic artery stenosis may share some risk factors. These findings emphasize the importance of better understanding the relationship between ophthalmic artery stenosis and age-related macular degeneration before endovascular therapy is pursued as a potential therapeutic option.

The current study has limitations. The retrospective design in patients who had undergone three-dimensional rotational angiography predominantly for the evaluation of intracranial aneurysms has the potential to introduce selection bias and limits the generalizability of the findings. We did not assess the interrater agreement of ophthalmic artery stenosis with less than fifty percent narrowing, nor did we grade the severity of stenosis in patients with at least moderate ophthalmic artery stenosis. The modest sample size limits the power of the analyses conducted. The observational nature precludes the inference of causality. Unmeasured and residual confounding may have contributed to the results.

In summary, we found that the assessment of ophthalmic artery stenosis on three-dimensional rotational angiography had excellent interrater agreement, suggesting that three-dimensional rotational angiography is a reasonable technique for diagnosing at least moderate ophthalmic artery stenosis prior to balloon angioplasty in future clinical trials.

Cigarette smoking and female sex were independently associated with ophthalmic artery stenosis, suggesting that age-related macular degeneration and ophthalmic artery stenosis may have some shared risk factors. Future research could evaluate visual function and retinal anatomy on optical coherence tomography and optical coherence tomography angiography in (visually) asymptomatic patients with ophthalmic artery stenosis to investigate whether there is a higher risk of early features of age-related macular degeneration.

## Data Availability

The data that support the findings of this study are available from the corresponding author upon reasonable request.

